# Beyond isolated cough events: AI-based tuberculosis screening through temporal analysis of cough sounds

**DOI:** 10.64898/2026.07.08.26357519

**Authors:** Ning Ma, Bahman Mirheidari, Guy J. Brown, Monde Muyoyeta, Nsala Sanjase, Minyoi M. Maimbolwa, Solomon Chifwamba, Seke Muzazu, Mary Kagujje

## Abstract

Tuberculosis (TB) is a major global health challenge, with many cases remaining undiagnosed due to limited access to screening and diagnostic services. Artificial intelligence (AI) systems based on cough sound analysis offer a scalable and accessible approach to TB screening, but most previous studies have analysed isolated cough events, despite the possibility that diagnostically useful information is encoded in the temporal dynamics of cough episodes. We evaluated an AI-based screening framework using cough recordings collected under real-world clinical conditions from 500 participants in Zambia, including 201 individuals with bacteriologically confirmed TB, 150 symptomatic patients with other respiratory diseases, and 149 healthy controls.

Using multiple pre-trained speech foundation models fine-tuned on cough sounds, we systematically investigated the influence of temporal context by varying the audio input window from 1 to 6 s, measured from the onset of each cough episode. Across all evaluated models, diagnostic performance consistently peaked with a 3 s input window, indicating that useful information extends beyond individual cough events and is encoded within the short-term temporal dynamics of cough episodes. The best audio-only model achieved an area under the receiver operating characteristic curve (AUROC) of 85.2% for distinguishing TB from all other participants and 80.1% for distinguishing TB from symptomatic non-TB respiratory disease. Incorporating demographic and clinical variables improved AUROC to 92.1% and 84.2%, respectively.

Performance remained robust across recording devices, participants with HIV co-infection, and varying acoustic conditions. These findings demonstrate that preserving temporal context improves AI-based cough screening for TB and suggest that analysing cough episodes, rather than isolated cough events, may enhance diagnostic performance in real-world settings. More broadly, the results highlight the importance of temporal context in the design of future respiratory sound datasets and AI-based diagnostic systems.

**Author summary:** Tuberculosis remains one of the world’s leading infectious diseases, and early detection is essential for reducing transmission and improving treatment outcomes. Cough is a common symptom of pulmonary tuberculosis and can be recorded using widely available devices such as smartphones, making cough sound analysis a potentially accessible screening approach. Most previous studies have analysed isolated cough sounds, but we investigated whether information contained in the temporal evolution of cough episodes could improve screening performance. Our study included people with bacteriologically confirmed tuberculosis, symptomatic patients with other respiratory diseases, and healthy individuals. Using AI models trained on cough sounds, we examined how much temporal context was needed for accurate screening. We found that analysing audio input window of approximately three seconds consistently outperformed shorter audio input, suggesting that diagnostically useful information is encoded not only within individual cough sounds but also in their short-term temporal dynamics. The system achieved strong performance in distinguishing tuberculosis from non-tuberculosis participants and remained robust across different recording devices and background acoustic conditions. These findings demonstrate the potential of AI-driven cough sound analysis as a practical and scalable screening tool for tuberculosis, particularly in resource-constrained settings, and highlight the importance of preserving temporal context when developing future respiratory sound datasets and diagnostic algorithms.

## Introduction

Tuberculosis (TB) remains a major global health challenge despite being preventable and curable. In 2024, there were an estimated 10.7 million new and relapse TB cases and 1.25 million deaths worldwide [1]. A significant proportion of TB-related mortality arises from undiagnosed and untreated disease, with nearly one quarter of all estimated TB cases remaining unrecognised [1]. Earlier detection through systematic screening is therefore essential to reduce transmission, improve treatment outcomes, and lessen the burden on health systems [2]. However, currently recommended screening approaches remain limited. Symptom-based screening has only moderate sensitivity and specificity [3], chest radiography requires equipment and infrastructure that are often unavailable in low-resource settings, and WHO recommends the use of C-reactive protein (CRP) testing and Molecular rapid diagnostic (mWRD) testing [4–6] only among people living with HIV (PLHIV) [7].

Artificial intelligence (AI) has the potential to improve TB screening by enhancing accessibility, efficiency, and diagnostic accuracy. AI-based approaches have already shown promise in medical imaging, disease detection, and physiological monitoring [8–15]. In 2021, the World Health Organization recommended AI-supported chest X-ray interpretation as an alternative to human readers for TB screening [2].

However, this approach still depends on imaging infrastructure and trained operators, which restricts deployment in many high-burden settings [5, 6]. These limitations motivate the development of alternative AI-enabled screening tools that rely on more accessible data sources, such as audio signals collected using low-cost microphones or smartphones.

Cough is a core symptom of pulmonary TB and may contain disease-related acoustic characteristics arising from inflammation and altered airway mechanics [16]. In clinical practice, cough quality, duration, and associated symptoms already inform diagnostic reasoning [17, 18]. AI-based cough analysis offers an opportunity to formalise and scale this process while reducing subjectivity [19]. Several studies have explored cough-based machine learning models for TB screening [16, 20–22], but progress has been limited by small datasets, inadequate representation of symptomatic non-TB participants, relatively simple machine learning approaches, and data collected under controlled acoustic conditions that may not reflect real-world deployment [16, 20–23]. More recently, large datasets such as CODA-TB [24] and advances in deep learning [25, 26] have accelerated development in this area.

Despite these advances, most cough-based AI systems analyse isolated cough events or very short audio segments centred on individual cough sounds, and temporal modelling of cough sounds remains largely unexplored. For example, the CODA-TB dataset primarily focuses on short cough events of approximately 0.5 s [24]. This implicitly assumes that diagnostically relevant information is contained within the acoustic characteristics of individual coughs. However, cough is a dynamic respiratory behaviour comprising inspiratory phases, cough bursts, inter-cough intervals, airflow recovery, and interactions between successive coughs rather than isolated events [27, 28], and objective cough monitoring increasingly recognises temporal cough patterns as clinically meaningful [29, 30]. We therefore hypothesised that disease-related information may be encoded not only in the acoustic properties of individual coughs but also in the temporal structure of cough episodes. The importance of such temporal context has received little attention in current TB cough datasets and benchmark studies. Whether longer audio input windows improve diagnostic discrimination, and how much temporal context is required, remains unknown.

In this study, we developed and evaluated a deep learning framework for TB screening using cough recordings collected under real-world clinical conditions in Zambia. Our primary objective was to investigate the role of temporal context in cough-based TB screening. Using multiple pre-trained speech foundation models, we systematically evaluated audio input durations ranging from one to six seconds to determine how temporal context influences diagnostic performance. We hypothesised that diagnostically useful information is not confined to isolated cough sounds but is also encoded within the temporal evolution of cough episodes. We further assessed whether the resulting models could discriminate between TB and non-TB participants under realistic recording conditions and remain robust across recording devices, participant subgroups, and potential acoustic confounding factors.

## Materials and methods

This cross-sectional diagnostic accuracy study followed the Standards for Reporting of Diagnostic Accuracy Studies (STARD) guidelines [31]. Ethical approval was obtained from the University of Zambia Biomedical Research Ethics Committee (approval number 3648-2023). Written informed consent was obtained from all participants before enrolment.

### Participants and data collection

Adults aged 18 years or older were recruited into three groups: (1) individuals with bacteriologically confirmed TB (TB^+^), defined by a positive Xpert MTB/RIF result; (2) symptomatic patients with respiratory disease but no TB (other respiratory diseases, OR); and (3) asymptomatic healthy controls (HC). The OR group was deliberately included because TB screening is typically performed among symptomatic individuals seeking care rather than healthy populations. This allowed assessment of the model’s ability to distinguish TB from clinically relevant alternative respiratory diagnoses and reduced the risk of overestimating performance through comparisons with healthy controls alone. Exclusion criteria for the TB^+^ group included previous TB, anti-TB treatment for more than three days, or a trace call result on Xpert MTB/RIF testing.

TB was excluded in the OR group using sputum Xpert testing and chest radiography. In the HC group, sputum Xpert MTB/RIF testing was performed. We aimed to recruit 550 participants (250 TB^+^, 150 OR, and 150 HC), exceeding the sample sizes of several previous studies of AI-driven acoustic analysis for TB screening [16, 20, 21, 23]. In total, 512 participants were enrolled between April 2023 and August 2024 from two Level-1 hospitals in Lusaka, Zambia: Kanyama and Chawama, both serving communities with a high burden of TB and HIV.

All participants underwent a brief clinical evaluation, including medical history and physical examination. TB^+^ participants were enrolled consecutively from TB clinics at the study sites. These individuals were referred from the same facility entry points as patients with other respiratory diseases. OR participants were recruited from symptomatic patients attending the same outpatient departments, and HC participants included asymptomatic caregivers and healthcare workers. To minimise demographic confounding, OR and HC participants were frequency-matched to the TB^+^ group by age and gender. Recruitment was balanced across sites and time periods to reduce potential site-specific and temporal biases.

Demographic and clinical information was recorded on paper forms and subsequently digitised in a secure electronic database. Cough recordings were collected in sound-attenuated outdoor Keter sheds using the same recording protocol across sites. The sheds were lined with foam to reduce acoustic reflections. The Kanyama shed was located approximately 100 m from road traffic near the chest clinic. The Chawama shed was similarly positioned near the rear hospital gate and adjacent to a church. A stereo microphone pair (RØDE M5) was positioned approximately 50 cm in front of the seated participant at head height. Simultaneous recordings were also captured using two Samsung Galaxy smartphones placed on the table in front of the participant to facilitate subsequent evaluation of device robustness and mobile deployment scenarios. A custom web-based application was used to synchronise and manage recordings across all devices. Audio files were saved temporarily on a laptop and later uploaded to secure cloud storage. Infection-control procedures included physical microphone barriers and ultraviolet disinfection between recording sessions.

Recording sessions were supervised by trained research staff. Participants were asked to produce at least three voluntary cough episodes, with each episode typically containing two to three individual cough events. Importantly, recordings were preserved as complete cough episodes rather than being segmented into isolated cough sounds at the point of acquisition. This design choice was motivated by the central hypothesis of the study that diagnostically useful information may be encoded not only around peak coughs but also in their short-term temporal dynamics. Preserving complete cough episodes enabled systematic investigation of how temporal context influences diagnostic performance.

### Audio preprocessing and cough segmentation

All recordings from both the RØDE microphones and the smartphones were downsampled to 16 kHz mono and trimmed to remove silence. Cough episodes were automatically extracted using an energy-based detector, with 200 ms of leading and trailing audio retained to preserve the acoustic context. Cough episodes boundaries were then manually reviewed and adjusted by human evaluators. Segmentation was performed using RØDE microphone audio only. Smartphone recordings were subsequently time-aligned using cross-correlation so that cough boundaries matched the reference episodes exactly. Each episode was labelled according to the participant’s diagnostic group: TB^+^, OR, or HC.

The final cough sound dataset comprised 3,117 cough episodes, with a total cough-only duration of 3 hours 56 minutes. Unlike datasets that isolate individual cough events, each cough episode may contain multiple coughs providing temporal information beyond individual cough sounds. The average cough episode duration was 4.55 s (range 2–9 s), with 87.8% of episodes shorter than 6 s. This distribution enabled systematic investigation of the size of the temporal window required for accurate TB screening.

Although recordings were collected in outdoor clinical environments with natural background noise, cough episodes had a high signal-to-noise ratio, averaging 30 dB. Signal-to-noise ratio was comparable across the two study sites (29.6 dB versus 31.9 dB).

### AI model development

The primary objective of this study was to investigate whether diagnostically useful information is encoded within the temporal dynamics of cough episodes. We therefore employed several pre-trained speech foundation models as a common modelling framework for analysing cough sounds across different temporal contexts (Fig. 1). The evaluated models included Wav2Vec2 [32], WavLM [33], HuBERT [34], Data2Vec [35], and Whisper [36]. These architectures generate contextualised temporal representations from raw audio waveforms and are therefore well suited to modelling acoustic dependencies that extend beyond individual cough events. Using multiple architectures also allowed us to determine whether any observed temporal-context effects were model-specific or represented a more general property of cough acoustics.

**Fig 1.**
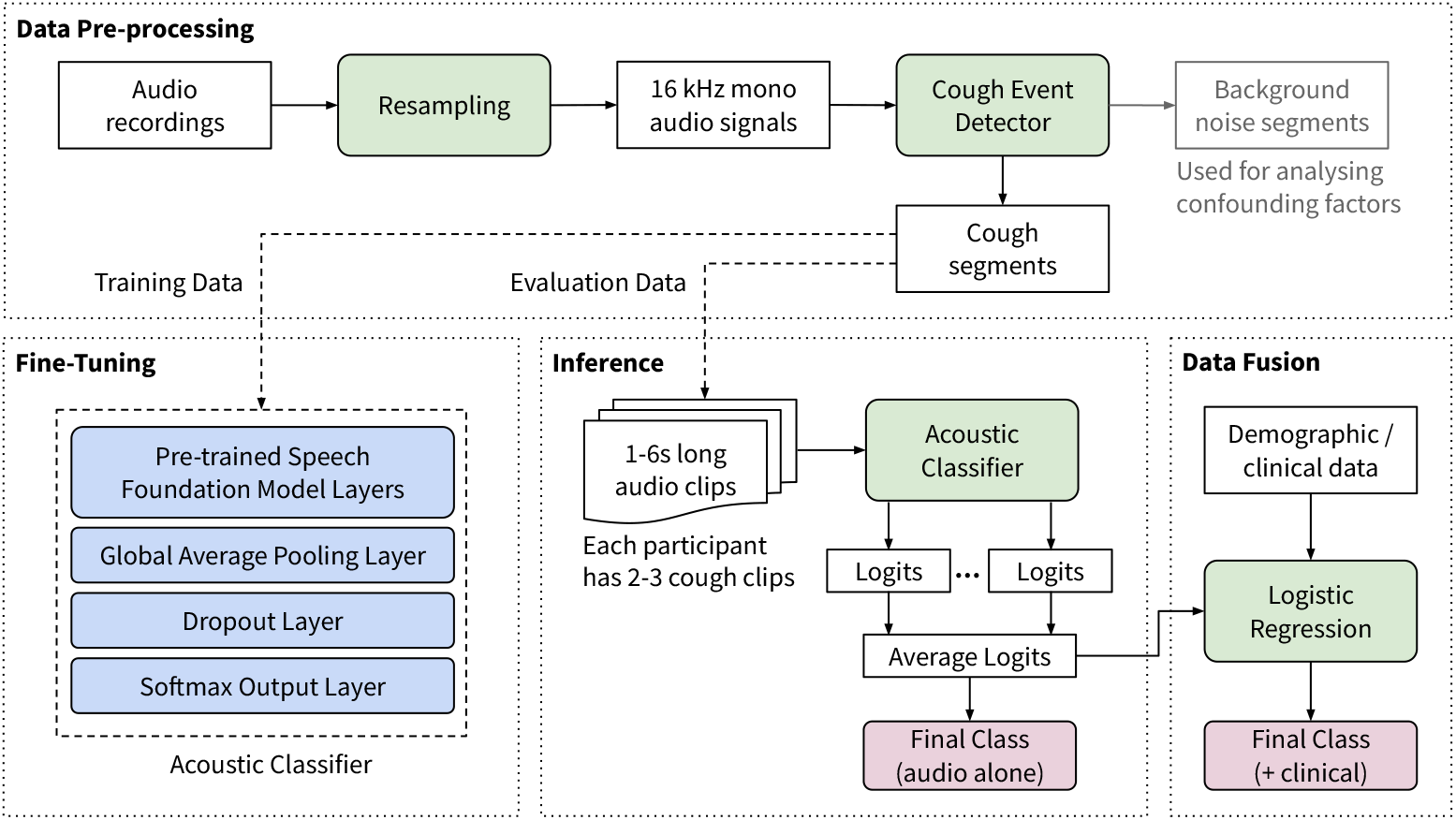
Pipeline used to investigate the role of temporal context in AI-based tuberculosis screening. Cough episodes were analysed using temporal windows ranging from one to six seconds to determine the amount of temporal information required for accurate classification.

For three-way classification (TB^+^, OR, HC), each foundation model was fine-tuned with an added classifier head comprising global average pooling, dropout (0.5), and a softmax output layer. Fine-tuning used 5 epochs, batch size 8, learning rate 3 × 10^*−*5^, warm-up ratio 0.1, and gradient accumulation over 8 steps. All experiments used stratified 10-fold cross-validation at the participant level. Cough episodes from a given participant were assigned exclusively to either the training or evaluation fold to prevent data leakage. Participant-level predictions were obtained by averaging episode-level softmax probabilities across all cough episodes from that participant and assigning the class with the highest mean probability.

Because demographic and clinical characteristics are known to be associated with TB risk, we also investigated whether cough acoustics provide additional diagnostic information beyond these routinely available variables. Audio-based predictions were therefore combined with demographic and clinical information using an ensemble stacking approach [37]. For each participant, softmax logits from the acoustic model were concatenated with age, gender, body mass index (BMI), and symptom variables to form a joint feature vector. A logistic regression (LR) meta-classifier was then trained to generate the final prediction. Training used both microphone and smartphone recordings to maximise variability, while evaluation was stratified by device type.

### Investigating the role of temporal context in cough episodes

The central aim of this study was to determine how much temporal context is required for effective cough-based TB screening. Previous studies have typically analysed isolated cough events or very short audio segments, assuming that diagnostically useful information is contained within individual cough sounds. To test this assumption, we systematically varied the duration of analysed audio while keeping all other aspects of the modelling pipeline unchanged. Audio input window was systematically varied from 1 to 6 s, measured from the onset of each cough episode (Fig. 2). When a cough episode exceeded the selected duration, it was truncated; when shorter, it was zero-padded.

**Fig 2.**
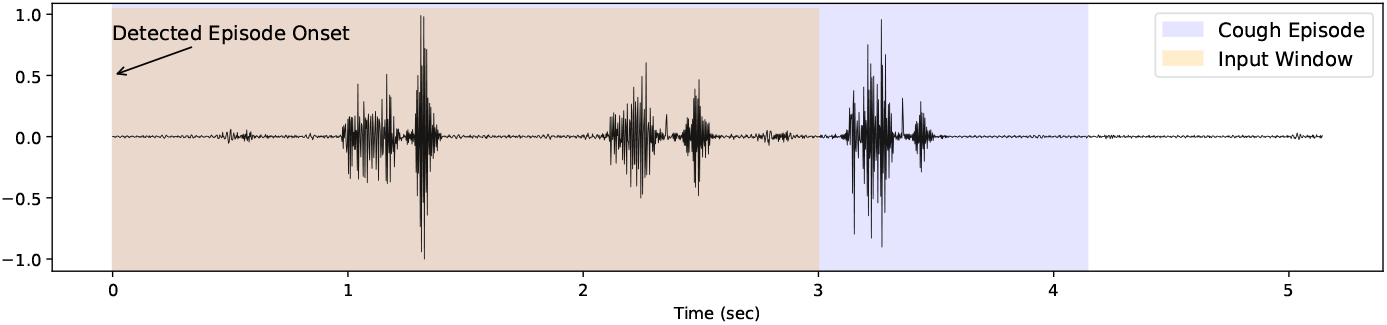
Illustration of the relationship between a cough episode and the analysed audio input window. A cough episode may comprise multiple cough events separated by short pauses. Audio input windows of varying duration (1–6 s) were extracted from the detected onset of each cough episode (illustrated here using the optimal 3 s window) and used as input to the AI models. The input window represents the temporal context available to the model for classification.

Shorter windows primarily capture isolated cough sounds, whereas longer windows increasingly incorporate inter-cough dynamics, inspiratory recovery phases, and surrounding respiratory context. By evaluating multiple temporal windows across several foundation-model architectures, we sought to determine whether diagnostically useful information is confined to individual cough events or distributed across the temporal evolution of cough episodes.

### Experimental design and evaluation

As a baseline, we trained a logistic regression model similar to [20] using mel-frequency cepstral coefficient (MFCC) features (40 coefficients plus delta and delta-delta). Model performance was evaluated using the area under the receiver operating characteristic curve (AUROC) across three binary diagnostic tasks: TB^+^ versus all other participants (TB^+^/Rest), TB^+^ versus symptomatic non-TB respiratory disease (TB^+^/OR), and TB^+^ versus healthy controls (TB^+^/HC). AUROC values were calculated by aggregating participant-level predictions across all held-out folds of the participant-level 10-fold cross-validation. Additional metrics included sensitivity, specificity, positive predictive value (PPV), negative predictive value (NPV), and F1-score.

The best-performing model on RØDE microphone recordings was selected for further analyses, including evaluation on participants with HIV co-infection, threshold-based screening analyses, and inference using smartphone recordings to assess robustness across recording devices and simulate mobile deployment scenarios. The effect of varying the classification threshold from 0.35 to 0.55 was examined. Confidence intervals for performance metrics were estimated using 10,000 bootstrap resamples of model predictions. Group differences in age were assessed using the Kruskal–Wallis test for three-group comparisons and the Mann–Whitney U test for two-group comparisons. Differences in gender distribution were assessed using the chi-square test.

To investigate the potential influence of acoustic confounding, trained audio-only models were additionally evaluated using non-cough background audio extracted from the same recordings. This analysis was designed to determine whether the model had learned to exploit characteristics of the acoustic background rather than cough acoustics.

## Results

### Characteristics of participants

The demographic and clinical characteristics of participants are summarised in Table 1. Of the 512 participants enrolled, 12 were excluded due to missing audio recordings, resulting in a final cohort of 500 participants, including 201 (40%) in the TB^+^ group, 150 (30%) in the OR group, and 149 (30%) in the HC group.

**Table 1.**
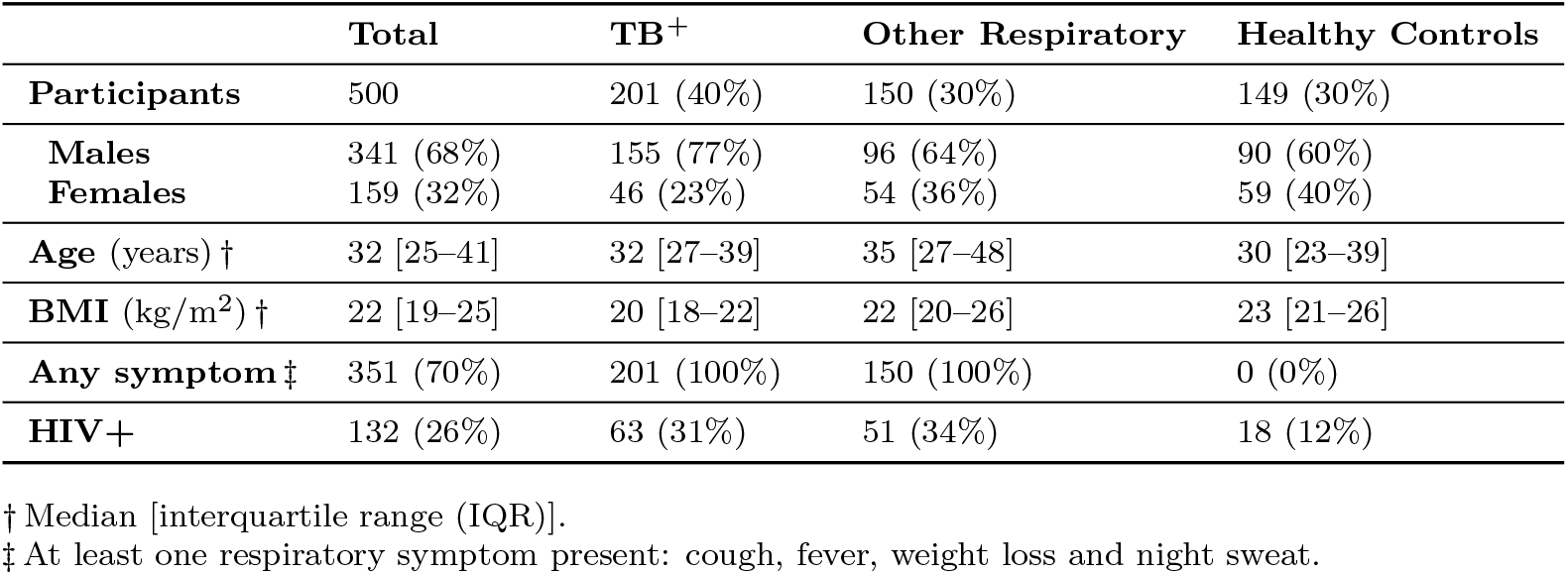
Demographic and clinical characteristics of study participants.

During recruitment, OR and HC participants were frequency-matched to the TB^+^ group by age and gender to reduce demographic confounding. However, as is common in real-world clinical studies, complete matching was not possible because participant enrolment reflected the characteristics of individuals presenting to healthcare services. Age differed across the three groups (Kruskal–Wallis test, *p <* 0.001), with OR participants tending to be older than both TB^+^ and HC participants. No significant age difference was observed between the TB^+^ group and the combined non-TB group.

Gender distribution also differed across groups (*χ*^2^ test, *p* = 0.002), with a higher proportion of males in the TB^+^ group (77%) than in the OR (64%) and HC (60%) groups. Median body mass index was lowest in the TB^+^ group (20 kg/m^2^ [IQR 18–22]), compared with the OR group (22 [IQR 20–26]) and HC group (23 [IQR 21–26]), consistent with the known association between active TB and weight loss. All participants in the TB^+^ and OR groups reported at least one respiratory symptom, whereas all HC participants were asymptomatic. Among the 500 participants, 132 (26.4%) were HIV-positive, including 63/201 (31%) in the TB^+^ group, 51/150 (34%) in the OR group, and 18/149 (12%) in the HC group.

### Temporal context is a key determinant of performance

Fig. 3 illustrates the relationship between temporal window size and screening performance for the TB^+^/Rest classification task, while Table 2 summarises AUROC values across all classifier architectures, input durations, and diagnostic tasks. A consistent pattern emerged across all evaluated foundation models: diagnostic performance improved as temporal context increased from 1 s to approximately 3 s before plateauing or declining. The highest AUROC values were generally observed with 3 s inputs, indicating that diagnostically useful information extends beyond isolated cough sounds and is encoded within the temporal dynamics of cough episodes.

**Table 2.** Comparison of AUROC with different classifier architectures and audio input durations. Fold-wise AUROC variability is not reported, as pooled AUROC was used to provide a single, stable estimate of diagnostic discrimination across the full cohort. Best results are in bold.

| Classifier | Model | Task | Audio Input Duration |  |  |  |  |  |
| --- | --- | --- | --- | --- | --- | --- | --- | --- |
| Architecture | Size |  | 1 s | 2 s | 3 s | 4 s | 5 s | 6 s |
| LR | 120 | TB<sup>+</sup>/ Rest | 76.4% | 79.6% | 78.8% | 75.3% | 75.7% | 70.2% |
|  |  | TB<sup>+</sup>/ OR | 71.5% | 75.2% | 74.3% | 70.9% | 72.6% | 65.2% |
| Whisper | 74M | TB<sup>+</sup>/ Rest | 81.9% | 82.8% | 83.8% | 82.6% | 83.5% | 83.7% |
|  |  | TB<sup>+</sup>/ OR | 76.4% | 76.7% | 78.3% | 76.8% | 77.3% | 77.8% |
| **Wav2Vec2** | 95M | TB<sup>+</sup>/ Rest | 81.8% | 83.9% | **85.2%** | 84.8% | 83.0% | 83.3% |
|  |  | TB<sup>+</sup>/ OR | 76.4% | 78.7% | **80.1%** | 79.5% | 76.9% | 78.0% |
| WavLM | 95M | TB<sup>+</sup>/ Rest | 76.4% | 83.5% | 84.6% | 82.5% | 84.2% | 82.7% |
|  |  | TB<sup>+</sup>/ OR | 70.7% | 77.2% | 78.8% | 75.9% | 78.2% | 75.8% |
| HuBERT | 95M | TB<sup>+</sup>/ Rest | 78.9% | 84.1% | 84.8% | 83.6% | 84.1% | 83.5% |
|  |  | TB<sup>+</sup>/ OR | 72.2% | 78.2% | 78.8% | 77.8% | 78.1% | 77.3% |
| Data2Vec | 95M | TB<sup>+</sup>/ Rest | 81.6% | 84.1% | 85.1% | 84.2% | 83.4% | 84.5% |
|  |  | TB<sup>+</sup>/ OR | 75.6% | 78.6% | 79.9% | 78.5% | 78.4% | 78.9% |

**Fig 3.**
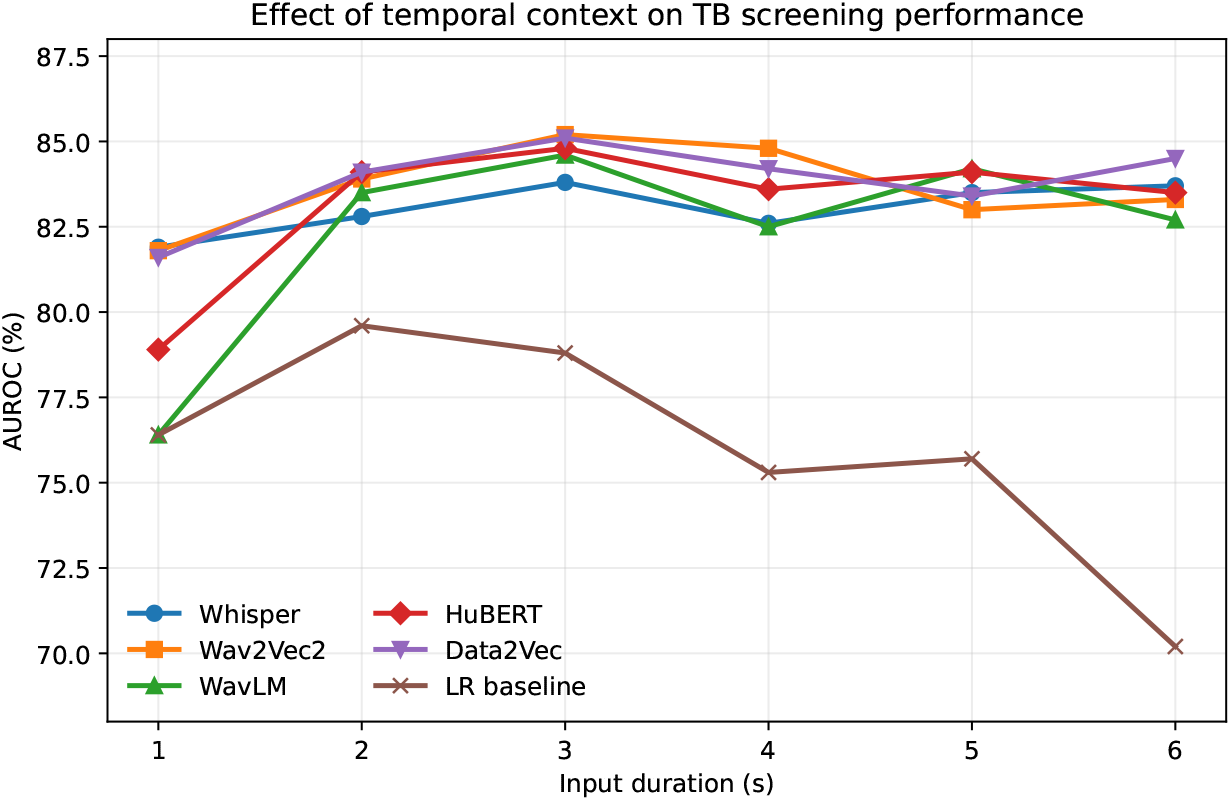
AUROC for the TB^+^/Rest classification task as a function of analysed temporal window size. Performance is shown for five speech foundation models and a logistic regression baseline. Across all foundation-model architectures, diagnostic performance consistently peaked at approximately 3 s, suggesting that diagnostically useful information extends beyond isolated cough sounds.

Across all three classification tasks, foundation model-based classifiers substantially outperformed the logistic regression baseline based on hand-crafted MFCC features.

The best overall performance was achieved by Wav2Vec2 using 3 s inputs, achieving AUROCs of 85.2% for TB^+^/Rest, and 80.1% for TB^+^/OR. Similar temporal trends were observed across Whisper, WavLM, HuBERT, and Data2Vec, suggesting that the observed benefit of temporal context was not specific to a single model architecture.

The influence of temporal context was particularly apparent when comparing 1 s and 3 s inputs. For example, Wav2Vec2 performance increased from 81.8% to 85.2% AUROC for TB^+^/Rest and from 76.4% to 80.1% for TB^+^/OR. Comparable improvements were observed for the other foundation models. In contrast, extending the analysed duration beyond 3 s did not consistently improve performance and, for some models, resulted in a modest decline. This finding suggests that a limited amount of temporal context is beneficial for TB screening, whereas longer windows may introduce additional variability without providing further diagnostically useful information. Overall, these results indicate that approximately 3 s of cough audio captures a favourable balance between isolated cough acoustics and broader cough-episode dynamics, supporting the hypothesis that temporal structure contributes meaningfully to AI-based TB screening.

### Complementary value of cough acoustics and clinical information

Using the best-performing Wav2Vec2 classifier with 3 s audio input, we next investigated whether demographic and clinical variables provided complementary information to cough acoustics. Specifically, we evaluated the incremental contribution of age, gender, BMI, and symptom variables using an ensemble stacking framework.

Performance is summarised in Table 3, and ROC curves for the main classification tasks are shown in Fig. 4.

**Table 3.** Comparison of classification performance using cough audio alone and in combination with demographic and clinical variables. Results are shown for the Wav2Vec2 classifier using a 3 s audio input window. Values in parentheses indicate the AUROC achieved using the corresponding demographic and clinical variables alone, without cough audio.

| Task | Features | AUROC | Sensitivity | Specificity | PPV | NPV | F1-score |
| --- | --- | --- | --- | --- | --- | --- | --- |
| TB<sup>+</sup> / Rest | Audio Alone | 85.2% | 73.2% | 82.3% | 73.5% | 82.0% | 73.3% |
|  | Audio + Gender | 85.7% (54.0%) | 74.1% | 83.9% | 75.6% | 82.8% | 74.8% |
|  | Audio + Age | 85.9% (49.6%) | 73.6% | 83.9% | 75.5% | 82.5% | 74.5% |
|  | Audio + BMI | 88.7% (76.5%) | 78.1% | 84.9% | 77.7% | 85.2% | 77.9% |
|  | Audio + Symptom | 90.1% (74.8%) | 80.6% | 83.2% | 76.4% | 86.4% | 78.4% |
|  | Audio + All Info | 92.1% (85.3%) | 81.1% | 85.9% | 79.5% | 87.1% | 80.3% |
| TB<sup>+</sup> / OR | Audio Alone | 80.1% | 73.2% | 72.0% | 77.8% | 66.7% | 75.4% |
|  | Audio + Gender | 80.4% (53.7%) | 74.2% | 76.0% | 80.6% | 68.7% | 77.2% |
|  | Audio + Age | 81.6% (55.1%) | 73.7% | 76.7% | 80.9% | 68.5% | 77.1% |
|  | Audio + BMI | 83.6% (72.3%) | 78.2% | 74.0% | 80.1% | 71.7% | 79.1% |
|  | Audio + Symptom | 80.3% (49.9%) | 80.6% | 66.7% | 76.4% | 72.0% | 78.4% |
|  | Audio + All Info | 84.2% (71.7%) | 81.1% | 72.0% | 79.5% | 73.9% | 80.3% |
| TB<sup>+</sup> / Rest (HIV+) | Audio Alone | 81.5% | 69.7% | 75.4% | 72.1% | 73.2% | 70.8% |
|  | Audio + All Info | 91.8% | 90.9% | 81.9% | 71.5% | 94.7% | 80.1% |

**Fig 4.**
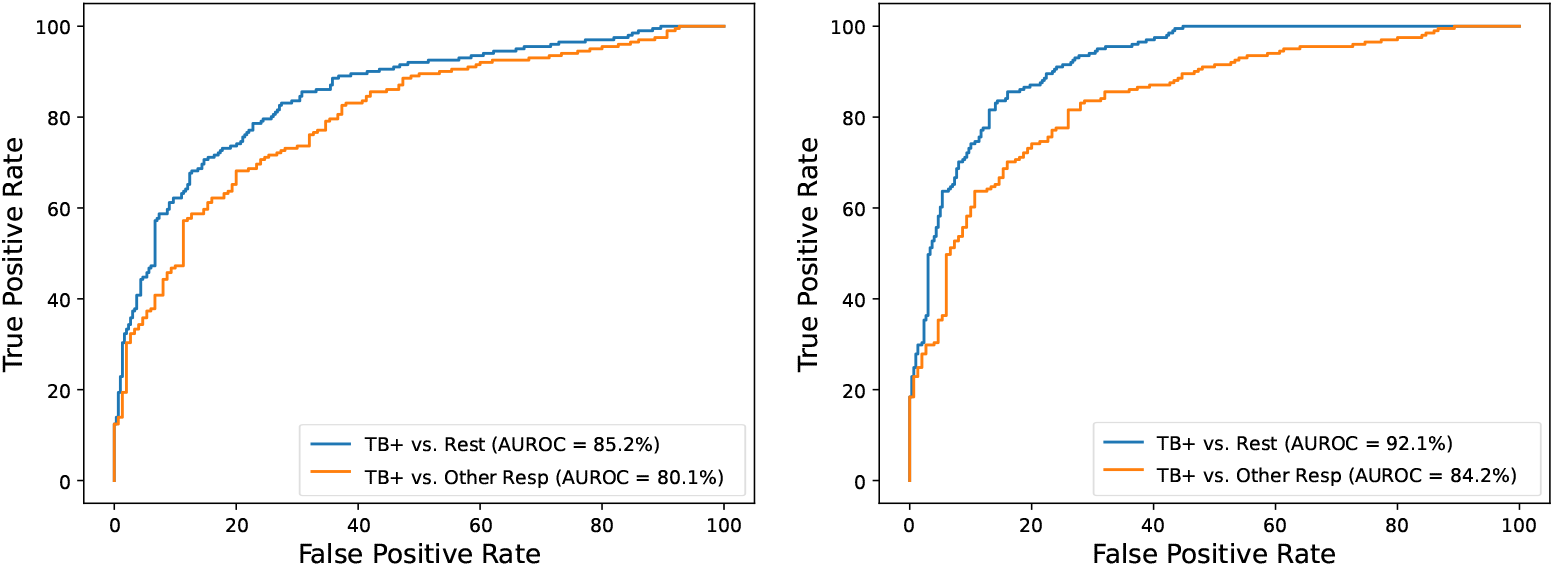
ROC curves for the Wav2Vec2 classifier using 3 s cough audio. Left: audio-only model for the TB^+^/Rest and TB^+^/OR tasks. Right: performance after combining cough acoustics with demographic and clinical information.

Using cough audio alone, the classifier achieved AUROCs of 85.2% for TB^+^/Rest and 80.1% for the more clinically challenging TB^+^/OR task. The lower performance for TB^+^/OR reflects the greater similarity between TB and other respiratory diseases, making this a more realistic and demanding screening scenario. Adding individual demographic or clinical variables resulted in modest performance improvements across tasks. The largest gains were observed when all variables were combined with cough acoustics. For TB^+^/Rest, AUROC increased from 85.2% to 92.1%, sensitivity increased from 73.2% to 81.1%, and F1-score increased from 73.3% to 80.3%. For TB^+^/OR, AUROC increased from 80.1% to 84.2%, while sensitivity increased from 73.2% to 81.1%.

To better understand the relative contributions of acoustic and non-acoustic information, Table 3 also reports the performance of models using demographic and clinical variables alone (AUROC values in parentheses). These variables achieved AUROCs of 85.3% for TB^+^/Rest and 71.7% for TB^+^/OR. Although demographic and clinical information was predictive, combining it with cough acoustics consistently improved performance. The benefit of cough audio was particularly evident for the clinically more challenging TB^+^/OR task, where AUROC increased from 71.7% using demographic and clinical variables alone to 84.2% when cough acoustics were incorporated. These findings suggest that cough sounds provide complementary information beyond routinely available demographic and clinical characteristics.

### Performance in participants with HIV co-infection

Because HIV co-infection is common among individuals undergoing TB investigation and may alter both clinical presentation and disease severity, we evaluated performance in the HIV-positive subgroup (*n* = 132). This subgroup comprised 63 participants with bacteriologically confirmed TB and 69 non-TB participants.

Using cough audio alone, the Wav2Vec2 classifier achieved an AUROC of 81.5%, with sensitivity of 69.7% and specificity of 75.4% (Table 3). Incorporating demographic and clinical information substantially improved performance, increasing AUROC to 91.8%, sensitivity to 90.9%, and negative predictive value to 94.7%. These findings suggest that cough acoustics remain informative in individuals living with HIV and that combining acoustic, demographic, and clinical information may be particularly beneficial in this clinically important population.

### Analysis of potential acoustic confounding

Because recordings were collected under real-world clinical conditions, we investigated whether model performance could be influenced by spurious acoustic cues unrelated to cough sounds. Although recruitment was balanced across sites and time periods, some day-wise imbalance remained. In particular, 48% of TB^+^ participants were recorded on days when only other TB^+^ participants were present, and a further 23% were recorded on days shared with OR participants. This raised the possibility that classifiers might inadvertently exploit environmental characteristics associated with specific recording sessions. In contrast, there was no significant difference in recording time across groups (one-way ANOVA, *F* (2, *N*− 3) = 1.59, *p* = 0.204).

Fig. 5 shows the long-term average spectra (LTAS) of cough episodes recorded using the RØDE microphone. The spectral profiles of TB^+^, OR, and HC coughs were broadly similar, with only minor differences across frequency bands, suggesting that group differences are not readily explained by simple average spectral characteristics.

**Fig 5.**
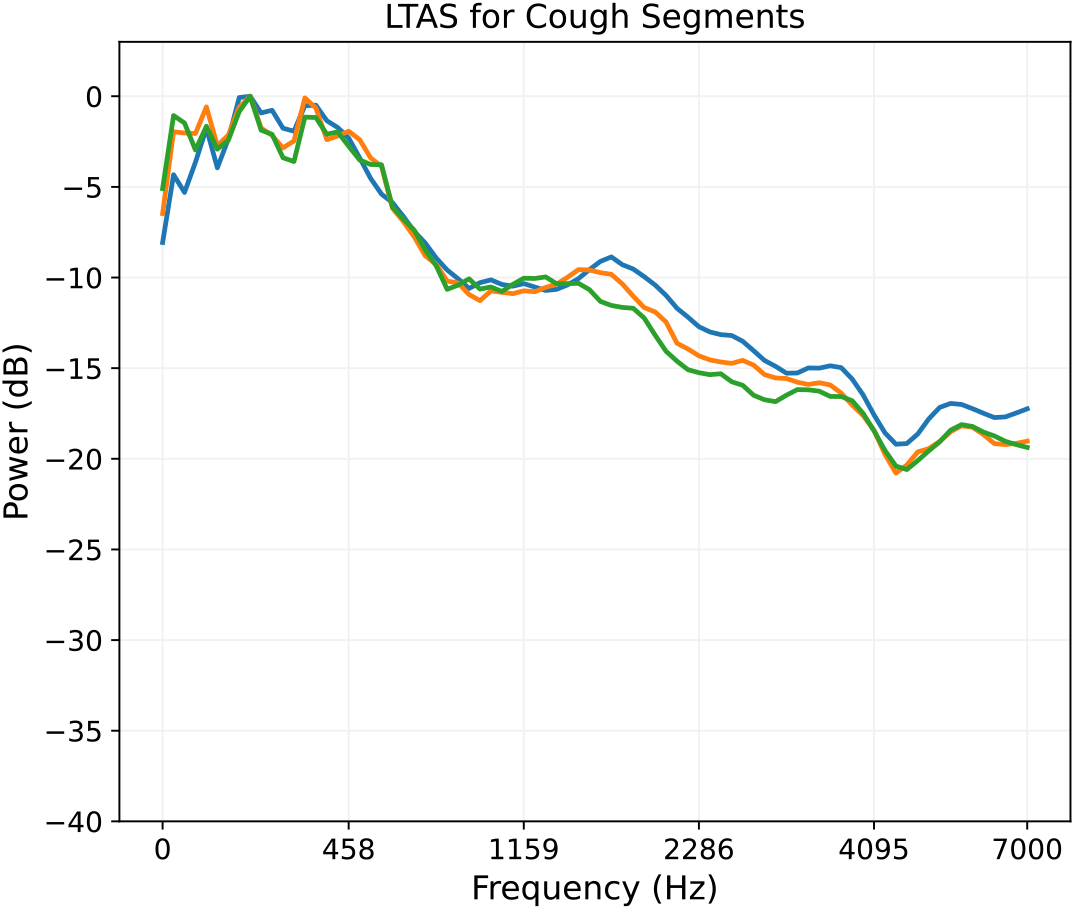
Long-term average spectra (LTAS) of cough audio episodes recorded using the RØDE M5 microphones. Average spectral profiles are broadly similar across TB^+^, OR, and HC groups, suggesting that group differences are not dominated by gross spectral characteristics.

To further assess the influence of environmental sounds, we performed an adversarial analysis using non-cough background audio extracted from the same recordings (Table 4). When evaluated on cough episodes, the model achieved an AUROC of 85.2% for the TB^+^/Rest task. In contrast, performance decreased to 58.6% when the same model was tested using background audio alone, approaching chance level. These findings suggest that the classifier primarily relied on disease-related cough characteristics rather than recording-specific environmental cues, providing additional evidence that the observed performance is unlikely to be explained by acoustic confounding.

**Table 4.** Effect of testing background using the proposed audio classifier.

| Task | Train Data | Test Data | AUROC |
| --- | --- | --- | --- |
| TB<sup>+</sup>/ Rest | Cough Sound | Background | 58.6% |
|  | Cough Sound | Cough Sound | **85.2%** |

### Screening performance across operating thresholds

The operating characteristics of the multimodal system (cough audio combined with all demographic and clinical variables) were evaluated across classification thresholds ranging from 0.35 to 0.55 (Table 5). As expected, increasing the classification threshold improved specificity at the expense of sensitivity. For the TB^+^/Rest task, a threshold of 0.35 prioritised case detection, achieving a sensitivity of 88.5% (95% CI 84.0–92.7) and a specificity of 77.9% (73.1–82.5). Increasing the threshold to 0.55 reduced sensitivity to 79.6% (73.9–85.0) while improving specificity to 87.9% (84.1–91.5). Negative predictive value remained high across all thresholds, ranging from 86.5% to 91.0%.

**Table 5.** Comparison of results for the proposed system (audio + all demographic/clinical information) with different thresholds. Bracketed figures show 95% confidence intervals based on 10,000 repeats.

| Task | Threshold | Sensitivity | Specificity | PPV | NPV | F1-score |
| --- | --- | --- | --- | --- | --- | --- |
| TB<sup>+</sup> / Rest | 0.35 | 88.5% (84.0–92.7) | 77.9% (73.1–82.5) | 72.9% (67.1–78.4) | 91.0% (87.3–94.4) | 79.9% (75.6–83.8) |
|  | 0.40 | 85.6% (80.5–90.2) | 78.9% (74.3–83.5) | 73.2% (67.5–78.9) | 89.1% (85.2–92.7) | 78.9% (74.5–83.0) |
|  | 0.45 | 82.6% (77.2–87.6) | 82.9% (78.6–87.1) | 76.5% (70.7–82.1) | 87.6% (83.6–91.3) | 79.4% (74.9–83.5) |
|  | 0.50 | 81.1% (75.5–86.3) | 85.9% (81.8–89.8) | 79.5% (73.9–85.0) | 87.1% (83.2–90.8) | 80.3% (75.8–84.3) |
|  | 0.55 | 79.6% (73.9–85.0) | 87.9% (84.1–91.5) | 81.6% (75.9–86.8) | 86.5% (82.6–90.3) | 80.5% (76.1–84.7) |
| TB<sup>+</sup> / OR | 0.35 | 88.5% (83.9–92.7) | 56.0% (48.0–63.8) | 72.9% (67.2–78.4) | 78.4% (70.4–86.0) | 79.9% (75.6–83.8) |
|  | 0.40 | 85.6% (80.6–90.3) | 58.0% (50.0–66.0) | 73.2% (67.4–78.9) | 75.0% (66.9–82.6) | 78.9% (74.6–82.9) |
|  | 0.45 | 82.6% (77.1–87.7) | 66.0% (58.3–73.5) | 76.5% (70.8–82.0) | 73.9% (66.2–81.2) | 79.4% (74.9–83.5) |
|  | 0.50 | 81.1% (75.5–86.4) | 72.0% (64.6–79.1) | 79.5% (74.0–84.8) | 73.9% (66.7–80.9) | 80.3% (76.0–84.3) |
|  | 0.55 | 79.6% (73.7–85.0) | 76.0% (68.9–82.6) | 81.6% (76.0–86.8) | 73.6% (66.5–80.4) | 80.5% (76.0–84.6) |

The TB^+^/OR task represented a more challenging and clinically realistic scenario because all participants presented with respiratory symptoms. At the lower threshold of 0.35, sensitivity was 88.5% (83.9–92.7), but specificity was limited to 56.0% (48.0–63.8). Increasing the threshold progressively improved specificity, reaching 76.0% (68.9–82.6) at a threshold of 0.55, while sensitivity decreased to 79.6% (73.7–85.0). Across thresholds, F1-scores remained relatively stable, ranging from 79.9% to 80.5%.

Overall, these results demonstrate the expected trade-off between sensitivity and specificity and indicate that threshold selection can be tailored to the intended screening context. Lower thresholds favour case detection and may be appropriate for triage applications, whereas higher thresholds reduce false-positive referrals at the cost of lower sensitivity.

### Performance on smartphone recordings

To assess the robustness of the proposed system to recording devices and simulate practical deployment scenarios, we evaluated the models using smartphone recordings acquired simultaneously with the RØDE microphone (Table 6). Overall, performance on smartphone recordings was comparable to that obtained using the RØDE microphone, demonstrating good generalisability across recording platforms. Using smartphone audio alone, the Wav2Vec2 classifier achieved an AUROC of 83.5% for the TB^+^/Rest task and 78.5% for the more clinically challenging TB^+^/OR task. Incorporating demographic and clinical information improved AUROC to 91.2% and 82.5%, respectively. For the TB^+^/Rest task, sensitivity increased from 72.6% to 80.6% and specificity from 79.9% to 84.3%. For TB^+^/OR, sensitivity similarly increased from 72.6% to 80.6%, while specificity decreased slightly from 70.7% to 68.7%. Compared with the corresponding RØDE microphone results, only modest reductions in performance were observed. These findings suggest that the proposed approach is robust to recording-device variability and support the feasibility of deploying cough-based TB screening using commodity smartphones in resource-constrained settings.

**Table 6.** Performance of the Wav2Vec2-based classifier on smartphone recordings. Results are shown for cough audio alone and for the multimodal system combining cough acoustics with demographic and clinical information.

| Task | Features | AUROC | Sensitivity | Specificity | PPV | NPV | F1-score |
| --- | --- | --- | --- | --- | --- | --- | --- |
| TB<sup>+</sup>/ Rest | Audio Alone | 83.5% | 72.6% | 79.9% | 70.9% | 81.3% | 71.7% |
|  | Audio + All Info | **91.2%** | **80.6%** | **84.3%** | **77.5%** | **86.6%** | **79.0%** |
| TB<sup>+</sup>/ OR | Audio Alone | 78.5% | 72.6% | **70.7%** | 76.8% | 65.8% | 68.1% |
|  | Audio + All Info | **82.5%** | **80.6%** | 68.7% | **77.5%** | **72.6%** | **79.0%** |

## Discussion

This study evaluated an AI framework for TB screening using cough audio recorded under real-world clinical conditions in Zambia. Across multiple speech foundation models, short but temporally informative 3 s input window consistently achieved the strongest performance. The best multimodal system, combining cough audio with demographic and clinical variables, showed strong discrimination between TB and non-TB participants and retained useful performance in the more clinically challenging comparison between TB and symptomatic non-TB respiratory disease. These findings support the potential of cough-based AI as a scalable and accessible screening approach in high-burden, resource-constrained settings.

A key finding of this study was the importance of temporal context in cough-based TB screening. Across all evaluated foundation models, performance consistently peaked at approximately 3 s, suggesting that diagnostically useful information extends beyond isolated cough sounds and is encoded within the short-term temporal dynamics of cough episodes. The consistency of this effect across model architectures suggests that it reflects properties of the cough signal rather than characteristics of any specific model. Shorter segments may fail to capture interactions between successive coughs and surrounding respiratory context, whereas longer segments may introduce additional variability and environmental noise. These findings have implications for current public datasets and benchmark studies, many of which focus on isolated cough events [24].

Future respiratory sound datasets should therefore consider preserving complete cough episodes and surrounding acoustic context.

Our findings also highlight the importance of including symptomatic non-TB participants when evaluating cough-based AI systems. Performance was consistently lower in the TB^+^/OR task than in the TB^+^/Rest task, which suggests distinguishing TB from other respiratory diseases is substantially more challenging than distinguishing TB from healthy controls. This distinction is clinically important because screening is typically performed among symptomatic individuals rather than healthy populations. By deliberately including a substantial OR group, our study provides a more realistic assessment of screening performance and reduces the risk of overly optimistic estimates reported in studies relying primarily on healthy controls [16, 20, 21].

The addition of demographic and clinical variables further improved performance. Although metadata-only models achieved reasonable discrimination, particularly for the TB^+^/Rest task, cough audio consistently provided complementary information, with the largest relative gains observed for the more clinically relevant TB^+^/OR comparison. These findings suggest that acoustic analysis and routinely available clinical information can be integrated to improve screening performance without requiring specialised measurements.

The system also demonstrated encouraging robustness across clinically important subgroups and recording devices. Among participants living with HIV, the multimodal model achieved an AUROC of 91.8%, although this subgroup was relatively small and the result should therefore be interpreted cautiously. HIV may influence respiratory physiology and symptom presentation [38], and future studies incorporating CD4 count, antiretroviral treatment status, and disease severity are needed to determine how well these findings generalise. Similarly, smartphone recordings achieved only modest reductions in performance compared with RØDE microphone recordings, supporting the feasibility of mobile and community-based deployment. Future work should investigate domain adaptation and device-aware training strategies to further improve robustness.

Potential acoustic confounding is an important concern in cough-based AI studies, especially when diagnostic groups are recorded under systematically different environmental conditions [39]. Although some day-wise imbalance remained in our dataset, targeted analyses showed that models trained on cough sounds performed close to chance when evaluated on background audio alone. Together with the absence of pronounced group differences in long-term average spectra, these findings suggest that the observed performance is unlikely to be explained primarily by recording-specific environmental cues. Nevertheless, continued attention to recording protocols, robustness testing, and external validation remains essential.

### Limitations and future directions

This study has several strengths, including the use of real-world recordings collected in a high-burden setting, deliberate inclusion of symptomatic non-TB participants, and systematic evaluation of temporal context, device variability, HIV subgroup performance, and potential acoustic confounding.

Nevertheless, several limitations should be acknowledged. First, participants were recruited from two sites within a single geographical region in Zambia, and broader validation across countries, languages, and recording environments is needed. Second, the relatively balanced cohort does not reflect TB prevalence in most screening populations, and predictive values may therefore differ in practice. Third, individuals with subclinical or previous TB were excluded, limiting conclusions regarding these clinically important groups. Fourth, although encouraging performance was observed among participants with HIV co-infection, more detailed immunological information was unavailable. Finally, while multiple foundation models were evaluated, computational efficiency and deployment constraints were not investigated in detail. An additional consideration is that direct comparison with previously published TB cough datasets and benchmark studies is challenging because many datasets contain only isolated cough events or very short audio segments. Consequently, it remains unclear whether existing approaches would achieve improved performance if longer cough episodes and richer temporal context were available.

Future work should focus on validation in diverse populations and prospective evaluation alongside established diagnostic pathways, including chest radiography and molecular testing. Preserving complete cough episodes in respiratory sound datasets will enable systematic evaluation of temporal context and fairer comparison between AI approaches. The importance of temporal context observed in this study suggests that analysing complete cough episodes, rather than isolated cough events, may improve the detection of TB and potentially other respiratory diseases.

## Conclusion

This study demonstrates that temporal context is an important component of cough-based AI screening for tuberculosis. Across multiple speech foundation models, approximately 3 s of cough audio consistently provided the strongest performance, indicating that diagnostically useful information extends beyond isolated cough sounds and is encoded within the short-term dynamics of cough episodes. Combining cough acoustics with simple demographic and clinical variables further improved discrimination, including in the more clinically relevant comparison between TB and other respiratory diseases. The proposed framework remained robust across HIV-positive participants, smartphone recordings, and potential acoustic confounding, supporting its potential for mobile and community-based screening. These findings highlight the importance of preserving temporal context in future respiratory sound datasets and AI systems and motivate prospective validation against existing diagnostic pathways in diverse populations.

## Data Availability

The TB cough sound dataset and associated code used in this study will be made available to qualified academic and non-profit researchers upon reasonable request following publication, subject to a data use agreement with the University of Sheffield and Centre for Infectious Disease Research in Zambia. Data access requests should be submitted to the corresponding author.

## Acknowledgements

This work was supported by the UK Higher Education Innovation Fund (X/179090) and the UK Engineering and Physical Sciences Research Council (EPSRC) Impact Acceleration Account (R/185787). The authors thank Ms Regina Banda and Ms Winfrida Mashili for their support with data collection, and Ms Sheba Nalwaba for assistance with data entry. We are also grateful to the management and staff of Kanyama and Chawama First-Level Hospitals for providing a supportive environment for data collection.

